# Impact of COVID-19 related lockdown on cognition and emotion: A pilot study

**DOI:** 10.1101/2020.06.30.20138446

**Authors:** Durjoy Lahiri, Souvik Dubey, Alfredo Ardila

## Abstract

COVID-19 pandemic has posed an unprecedented challenge in front of the world contributed mostly by social distancing and lockdown. Among several other effects this pandemic has wreaked havoc on the psychology and cognition of people across the globe. In this paper we attempted to find out the impact of lockdown and social isolation on the cognition and emotion of young healthy adults with high education (n=43) by means of a questionnaire sent through email. We found that more than 50% of the participants had some kind of emotional or cognitive (dysexecutive) symptoms, as calculated through emotional symptom index (ESI) and cognitive symptom index (CSI). The correlation between cognitive and emotional symptoms was also found to be moderately strong (0.59). Although it is a pilot study and larger samples are required to draw firm conclusion, the results argue in favor of a negative impact on the cognition and emotion of healthy educated young people caused by the COVID-19 related lockdown. It can be conjectured that, if taking an older sample with a lower education, emotional and cognitive changes would be more evident.

## Introduction

The pandemic of COVID-19 has posed an unprecedented challenge in front of the world. While several studies are devoted towards examining the clinical effects of SARS-CoV-2 infection, few published so far have investigated the psychological consequences of such a rare global health emergency (e.g., Dai et al, 2020; Wang et al, 2020). It can be anticipated, however, that toward a near future, a myriad of studies dealing with this topic will become available.Our experience from previous pandemics of MERS and SARS shows that during times of such large-scale emergency the non-infected population may suffer from several psychological adverse effects. Among these ill-effects on mental health, post-traumatic stress disorder (PTSD) is the forerunner followed closely by depression, anxiety, and other behavioral &psychological disorders (Jung et al, 2020; Park&Park, 2020; Wu, Chan & Ma, 2005).

Psychological symptoms are known to affect cognitive functioning particularly in the domain of attention and executive function. In this context two large meta-analyses may be referred to. Zakzanis, Leach & Kaplan (1998) found that specific components of executive functioning, namely verbal fluency and inhibition, are vulnerable in depression. By the same token, Snyder’s (2013) meta-analysis found, major depressive disorders associatedwith significant impairments on all measures of executive function.

A recent paper highlighting the neurobiological consequences of social isolation points out that remaining socially isolated can have deleterious effect on memory and executive dysfunction in the long run (Bzdok& Dunbar, 2020). It is understood that different sections of the population have different demands of executive function and hence the effects on this cognitive attribute will also vary. To our knowledge, no study thus far has analyzed the cognitive effects on general population consequent to the ongoing pandemic as well as the unprecedented measures to curb it, for instance social distancing and prolonged lockdown.

Education is known to be associated with increased cognitive reserve (Cammano-Isoma, 2006; Meng& D’Arcy, 2012). Prospective studies have shown that this association is valid in terms of level of cognitive functioning rather than the rate of cognitive decline (Wilson et al, 2009; Winnock et al, 2002). In other words, higher educated persons are not immune to the usual age-related cognitive decline (Van Dijk et al, 2008). In addition, higher cognitive reserve is thought to alter the clinical trajectories of dementia onset by delaying the symptom onset. In sum, there is evidence to believe that higher educated persons have better cognitive reserve and therefore may be better capable of coping with emergency situations such as the present one that puts the cognitive system under stress owing to the psychological effects of isolation. Similarly, young people are thought to have better cognitive reserve given the several mechanisms that influence cognitive decline with increasing age (Harada et al, 2013). Considering the aforementioned arguments, we believe that studying the cognitive effects of isolation on young and middle age educated individuals may be more sensitive as a screening investigation in this context.

In this background, we set out to explore the emotional and cognitive effects of isolation on adults with high level of formal education. The objective of the present study was to find out the prevalence of emotional and cognitive symptoms in the backdrop of COVID-19 lockdown among the educated persons. Besides, we also aimed to have a basic measure of the psychological symptoms prevalent in the same population. Here we report the preliminary findings of the pilot study that we conducted over last couple of months. We intend to take the present study forward with larger sample so as to have a deeper understanding of the problem.

## Methods

Self-reported questionnaire was developed focusing on symptoms related to executive dysfunction and emotional symptoms such as low mood and anxiety along with duration of the respective symptoms. The questionnaire is included in the Appendix. The questionnaire was administered in English language. Institutional ethics committee waived off supervision for this online survey.

Participants were young and middle age adults in the age range of 25 to 40 years with high education and without any pre-existing psychiatric or cognitive illness. Any person known to have been infected with SARS-CoV-2 or exposed to it was excluded from the tally. Potential participants fulfilling the inclusion criteria were requested through email to fill up the questionnaire. Among the respondents consecutive 43 were included in the study of which 39 responses were finally analyzed.

Basic statistical analysis was carried out calculating the percentage of different self-reported symptoms in the aforementioned domains. Indexation was applied taking into account the self-reported symptoms. Emotional symptom index (ESI) was computed based on the indices of the 3 questions related to low mood, anxiety, and sleep duration, while cognitive symptom index (CSI) was computed based on the symptoms related to apparent executive dysfunction, namely losing track of conversation, losing track of day and date, misplacing common objects of daily use, and increase in retrieval or processing time of information. Correlation co-efficient between the two composite indices, ESI and CSI was calculated to establish their inter-relationship.

### Statistical analysis

We have analyzed a sample of 39 respondents through ESI and CSI. Our parameter selection was normally distributed. Each single parameter is unique, mutually exclusive and distinct. Therefore an indexation technique was used to homogenize the entire classification. It consists of two simultaneous stages- (1) Indexation: by using the statistical technique of normalization (Actual-Minimum/Maximum-Minimum, value ranges from 0 to 1) ; (2) Weightage: by assigning equal weightage for all the indices. Hence ESI = 1/3(Low mood index+ Anxiety index+ Sleep index) and CSI = ¼ (Losing track of conversation index+ Losing track of day and date index+ Object misplacement index+ Index of processing/ retrieval). By correlating ESI and CSI we get a dimension of directional measure (r_xy_= Cov (X,Y)/ SD_x_, SD_y_).

## Results

A total of 43 responses were gathered from the online survey conducted for this study. Of these, 4 responses were excluded because they were incomplete and therefore 39 were included in the final analysis. Table 1 presents the characteristics of the sample. Eight participants reported pre-existing co-morbidities of which 5 had hypothyroidism (on replacement) and 4 reported hypertension (on medication). None of the participants reported any pre-existing psychiatric illness or symptoms related to cognitive difficulty.

**Table 1:**
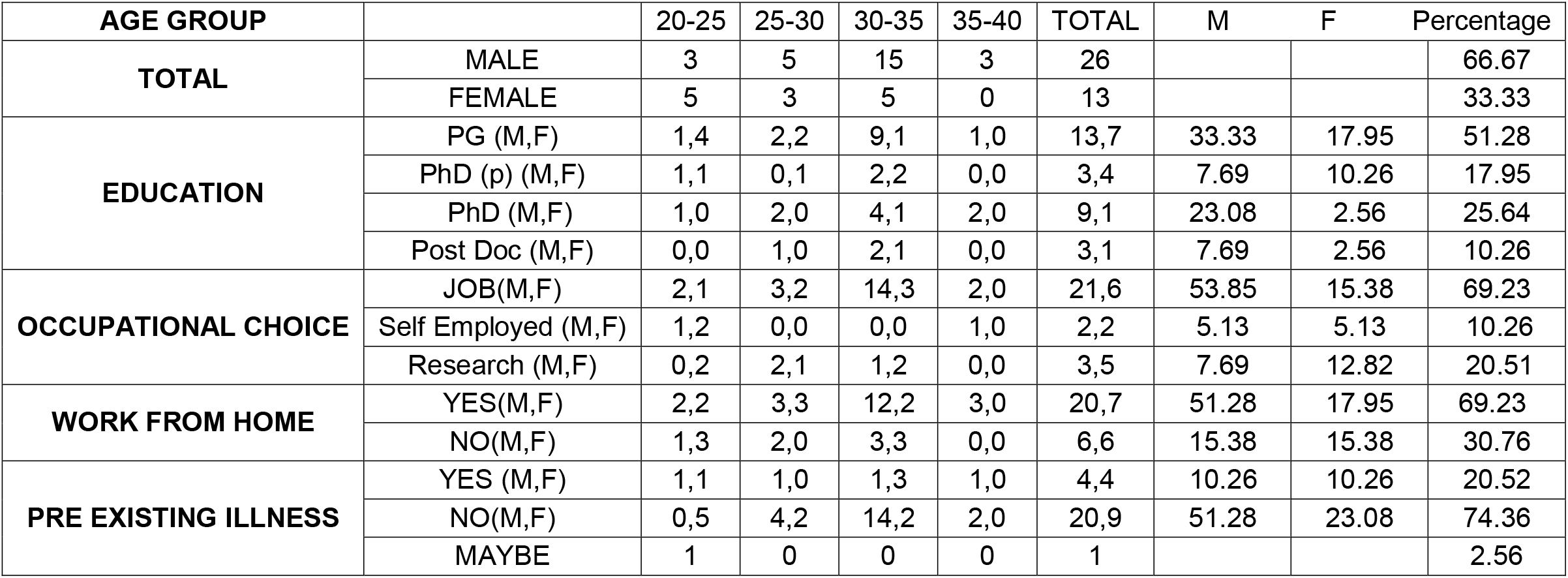
Demographic data; PhD (p): PhD pursuing; Educational qualification & minimum equivalent years of formal education: P.G.-18 years, PhD (p)-20 years, PhD-25 years, Post Doc-27 years

Table 2 presents the frequency of responses to the questions and Table 3 displays the calculated indices (ESI and CSI).

**Table 2:** Frequency of responses; Responses to the question of sleep duration were as follows: more than 6 hours-71.79%; 5 to 6 hours-10.25%; 4 to 5 hours-10.25%; <4 hours-5.12%

| Questions | Yes | No | May be |
| --- | --- | --- | --- |
| Anxious | 66.66 | 17.94 | 15.94 |
| Low mood | 76.92 | 23.07 | - |
| Misplacing common objects | 33.33 | 58.97 | 7.70 |
| Losing track of conversation | 23.07 | 69.23 | 7.70 |
| Losing track of day and date | 46.15 | 41.02 | 12.83 |
| Increase in retrieval /processing time | 33.33 | 61.53 | 5.14 |

**Table 3:** ESI and CSI

| ESI |  |  | CSI |  |  |  |
| --- | --- | --- | --- | --- | --- | --- |
| Low mood | Anxiety | Sleep | Losing track of conversation | Track of day and date | Misplacement of object | Retrieval or process |
| 0.53 | 0.33 | 0.70 | 0.71 | 0.57 | 0.72 | 0.57 |
| 0.52 |  |  | 0.64 |  |  |  |

Low mood index was found to be 0.52; that means, 52% in average presented emotional changes; while anxiety index was 0.33 and sleep index was 0.70. The composite index of these 3 indices, termed as ESI, was 0.52. Similarly, indices for conversational track, date and day track, object misplacement and retrieval/processing of information were observed to be 0.70, 0.56, 0.72, 0.57 respectively and the composite index, termed as CSI, was found to be 0.33. When correlation between ESI and CSI were computed, the value came out to be 0.597, suggestive of a moderately strongcorrelation between the two signifying that ESI and CSI can be co-operative through proper measure.

## Discussion

Although this is a pilot study with a limited sample and a small amount of questions, it is evident that an important percentage of the participants reported some emotional and cognitive changes associated with the lockdown situation that they are living in. About 50% of the participants are feeling as having some emotional and cognitive changes, but without any specific quantification of their magnitude.

Our participants presented two important demographic characteristics: they were young or middle age adults on one hand, and they had a high level of education on the other. It can be conjectured that, if taking an older sample with a lower education, emotional and cognitive changes would be more evident. There is a significant association between the sensitivity to stressful situations and age; and also coping strategies in stressful situations are associated with age (De Minzi&Sacchi, 2005). Education, on the other hand, is considered critical in cognitive performance, not only in young adults, but even in elders (e.g., Wilson et al. 2019). Nonetheless, within the current conditions, about 50% of the participants reported emotional and cognitive changes associated with the lockdown situation currently existing in a significant number of countries world-wide. At this point it is impossible to know if these changes are going to have long-lasting consequences, once the lockdown is over (Greenberg, Carr& Summers, 2002) but it is known that there are individual differences in the vulnerability to stress (Kessler, 1979). Thus, not everyone responds in the same way to the stress associated with the lockdown and probably, long-term consequences may be different.

Cognition and emotion are thought to affect each other. Together they form the basic building blocks of metacognition (Akturk & Sahin, 2011) and play significant role in orchestrating theory of mind (Airenti, 2015). Cognition, emotion, motivation, planning, organization, prudence and related executive abilities together with psyche also form the basis of stress coping. COVID-19 pandemic has unraveled difficulties in stress coping with its psychiatric manifestations. Stress is known to have direct association with dysfunctional pre-frontal network of cognitive circuit (Selemon, Young, Cruz & Williamson, 2019; Soares et al, 2013) without completely defying the contributory effect of depression and low mood. The present study revealed attention deficit and dysexecution related to lockdown.

There is a diversity of limitation is this study related not only to the questionnaire that was used, the characteristics of the sample, and the limited number of participants, but also with the specific city and country where the sample was collected. The severity of the COVID-19 pandemic and social response to it, has been highly variable across regions and countries (Hopman, Allegranzi&Mehtar, 2020). Generalization of current results should be extremely cautious.

## Data Availability

Data will not be available as it is a pilot study and we are expanding the sample.

## Acknowledgements

We are sincerely thankful to Saptarshi Chowdhury (Assistant Professor, Post doctoral fellow, Calcutta University, India) for his immense support during collection and analysis of data for this pilot study.

## QUESTIONNAIRE: 1

1 NAME:

2. AGE:

3. CONTACT NO:

4. GENDER:

5. EDUCATIONAL QUALIFICATION:

6. OCCUPATION:

7. PRESENTLY WORKING FROM HOME: DURATION:

8. PRE EXISTING ILLNESS:

9. PREVIOUSLY USED MEDICATION:

10. DO YOU LACK TRACK OF CONVERSATION? DURATION

11. DO YOU FIND ANY DIFFICULTY TO TRACK DAY & DATE? DURATION

12. DO YOU MISPLACE COMMON OBJECT OF DAILY USE? DURATION

13. DO YOU REQUIRE ANY INCREASED TIME TO RETRIEVE INFORMATION? DURATION

14. DO YOU FEEL ANXIOUS ABOUT LOCKDOWN? DURATION

15. ARE YOU IN A LOW MODD DURING LOCK DOWN? DURATION

16. HOURS OF SLEEP AT NIGHT : DURATION

## References

Airenti, G. (2015). Theory of mind : a new perspective on the puzzle of belief ascription. Frontiers in Psychology, 6, 1184.

Arktuk, A.O., Sahin, I. (2011). Literature review on metacognition and its measurement. Procedia Social and Behavioral Sciences, 15, 3731–3736.

Bzdok, D. & Dunbar, R.I.M. (2020). The Neurobiology of Social Distance.Trends in Cognitive Sciences. doi:https://doi.org/10.1016/j.tics.2020.05.016

Caamano-Isoma, F., Corral, M., Montes-Martinez, A., Takkouche, B. (2006). Education anddementia: a meta-analytic study. Neuroepidemiology, 26,226–232.

Dai, Y., Hu, G.,Xiong, H., Qiu, H. & Yuan, X. (2020). Psychological impact of the coronavirus disease 2019 (COVID-19) outbreak on healthcare workers in China. medrxiv. https://doi.org/10.1101/2020.03.03.20030874

De Minzi, M. C. R., & Sacchi, C. (2005). Stressful situations and coping strategies in relation to age. Psychological Reports, 97(2), 405–418.

Greenberg, N., Carr, J. A., & Summers, C. H. (2002). Causes and consequences of stress. Integrative and Comparative Biology, 42(3), 508–516.

Harada, C.N., Marissa, C., Love, N. & Triebel, K. (2013). Normal cognitive ageing. Clinical Geriatric Medicine, 29(4), 737–752. doi:10.1016/j.cger.2013.07.002.

Hopman, J., Allegranzi, B., & Mehtar, S. (2020). Managing COVID-19 in low-and middle-income countries. Jama, 323(16), 1549–1550.

Jung, S.J. & Jun, J.Y. (2020). Mental Health and Psychological Intervention AmidCOVID-19 Outbreak: Perspectives from South Korea.Yonsei Medical Journal, 61(4),271–272 https://doi.org/10.3349/ymj.2020.61.4.271

Kessler, R. C. (1979). A strategy for studying differential vulnerability to the psychological consequences of stress. Journal of Health and Social Behavior, 100–108.

Meng, X., D’Arcy, C. (2012). Education and dementia in the context of the cognitive reservehypothesis: a systematic review with meta-analyses and qualitative analyses. PLoS One, 7,e38268.

Park, S. C., & Park, Y. C. (2020). Mental Health Care Measures in Response to the 2019 Novel Coronavirus Outbreak in Korea. Psychiatry investigation, 17(2), 85–86. https://doi.org/10.30773/pi.2020.0058

Selemon, L. D., Young, K. A., Cruz, D. A., & Williamson, D. E. (2019). Frontal Lobe Circuitry in Posttraumatic Stress Disorder. Chronic stress (Thousand Oaks, Calif.), 3, 2470547019850166. https://doi.org/10.1177/2470547019850166

Soares, J.M., Sampaio, A., Ferreira, L.M., Santos, N.C., Marques, P……. Sousa, N. (2013). Stress impact on resting state brain networks. Plos One, 8 (6), e 0066500

Snyder, H. R. (2013).Major depressive disorder is associated with broad impairments on neuropsychologicalmeasures of executive function: A meta-analysis and review. Psychological Bulletin,139(1), 81–132. https://doi.org/10.1037/a0028727.

Van Dijk, K.R.A., Van Gerven, P.W.M., Van Boxtel, M.P.J., Van der Elst, W. & Jolles, J. (2008). No protective effects of education during normal cognitive aging: results from the 6-year follow-up of the Maastricht Aging Study. Psychol Aging, 23, 119–130.

Wang, H.,Xia, Q., Xiong, Z.,Li, Z., Xiang, W., Yuan, Y., Liu, Y., Li Z. (2020). The psychological distress and coping 1 styles in the early stages of the 2019 coronavirus disease (COVID-19) epidemic in the general mainland Chinese population: a web-based survey. medrxiv. https://doi.org/10.1101/2020.03.27.20045807

Wilson, R. S., Yu, L., Lamar, M., Schneider, J. A., Boyle, P. A., & Bennett, D. A. (2019).Education and cognitive reserve in old age. Neurology, 92(10), e1041–e1050.

Wilson, R.S., Hebert, L.E., Scherr, P.A., Barnes, L.L., Mendes de Leon, C.F. & Evans, D.A. (2009). Educational attainment and cognitive decline in old age. Neurology, 72, 460–465.

Wilson, R.S., Yu, L., Lamar, M., Schneider, J.A., Boyle, P.A. & Bennett, D.A. (2019). Education and cognitive reserve in old age. Neurology, 92, e1041–e1050. doi:10.1212/WNL.0000000000007

Winnock, M., Lelenneur, L., Jacqmin-Gadda, H., Dallongeville, J., Amouyd, P. & Dartigues, J.F. (2002). Longitudinal analysis of the effect of apolipoprotein E e4 and education oncognitive performance in elderly subjects: the PAQUID study. Journal of Neurology Neurosurgery and Psychiatry,72,794–797.

Zakzanis, K.K., Leach, L.,& Kaplan, E. (1998).On the nature and pattern of neurocognitive functionin major depressive disorder. Neuropsychiatry, Neuropsychology, and Behavioral Neurology,11(3), 111–119.

